# Heterogeneous effects of physical activity on physiological stress during pregnancy

**DOI:** 10.1101/2025.03.30.25324909

**Authors:** Jenifer Rim, Qi Xu, Xiwei Tang, Tamara Jimah, Yuqing Guo, Annie Qu

## Abstract

Pregnancy is a critical period characterized by profound physiological and psychological adaptations that can significantly impact both maternal and fetal health outcomes. Thus, it is imperative to implement targeted and evidence-based interventions to enhance maternal well-being during the prenatal period. Mobile health (mHealth) technologies enable continuous, real-time monitoring of both physiological and psychological states, providing detailed insights into health behaviors and individual responses in natural settings. This study leveraged mHealth technologies, including the Oura smart ring and ecological momentary assessment (EMA) via a mobile app, to examine how emotional distress influences the relationship between physical activity (PA) and heart rate variability (HRV), an indicator of physiological stress during pregnancy. Consenting participants, aged 18-40 years, with a healthy singleton pregnancy in the second trimester, were enrolled in the study. Our findings revealed that among participants experiencing emotional distress, increased PA was associated with higher HRV, indicating lower physiological stress. In particular, this beneficial effect was more pronounced on days with elevated emotional distress. These findings suggest that engaging in physical activity may help protect pregnant women against autonomic dysregulation associated with emotional distress, potentially supporting maternal cardiovascular health. By utilizing mHealth technologies for real-time data collection, our study highlights the potential for personalized and adaptive interventions that promote maternal well-being by encouraging physical activity as a strategy to mitigate the physiological effects of emotional distress during pregnancy.

**Author summary:** Pregnancy involves significant physiological and psychological changes that impact maternal and fetal health. Heart rate variability (HRV), a key biomarker of autonomic function and cardiovascular health, reflects the body’s ability to regulate physiological stress. In this study, we used mobile health (mHealth) technologies, including the Oura smart ring and ecological momentary assessment (EMA) through a mobile app, to examine how emotional distress influences the relationship between physical activity (PA) and HRV during pregnancy. Our findings indicate that PA protects against autonomic dysregulation linked to emotional distress. On days with emotional distress, engaging in more PA was associated with higher HRV, indicating lower physiological stress. This positive effect was even stronger on days with increased emotional distress. By leveraging real-time monitoring, our study highlights the value of mHealth in capturing dynamic interactions between emotional and physiological states. These insights demonstrate the potential for personalized interventions using mHealth technologies to support maternal well-being by encouraging PA as a strategy to better manage physiological stress during pregnancy.

## 1 Introduction

Pregnancy is a critical period characterized by profound physiological and psychological adaptations that can significantly impact both maternal and fetal health outcomes. Among these, physiological stress, defined as the body’s autonomic and endocrine responses to perceived challenges, constitutes a crucial determinant of pregnancy health [1]. The impact of physiological stress on maternal health and fetal development is well-studied [2, 3, 4]. In particular, excessive physiological stress during pregnancy has been linked to adverse outcomes such as preterm birth, low birth weight, and developmental issues in the child [5, 3, 6], as well as an increased risk of maternal conditions such as hypertension, gestational diabetes, and postpartum depression [7]. Given these consequences, understanding factors that influence physiological stress regulation during pregnancy is essential for informing interventions that promote maternal and fetal health.

Among the various physiological markers to assess physiological stress, heart rate variability (HRV) constitutes a well-established indicator of autonomic nervous system function and cardiovascular health regulation [8]. HRV refers to the variation in time intervals between heartbeats and reflects the balance between sympathetic (stress-related) and parasympathetic (relaxation-related) nervous system activity [9]. Higher HRV is generally associated with good cardiovascular health and resilience to stress, whereas reduced HRV indicates compromised autonomic regulation and increased physiological stress exposure [10]. Thus, HRV not only reflects physiological stress resilience but also serves as an indicator of cardiovascular fitness, both of which are important for understanding and informing pregnancy health. This is particularly relevant during the second trimester, as this period is marked by significant cardiovascular and physiological adaptations [11, 12].

Advancements in mobile health (mHealth) technologies have enabled continuous monitoring of HRV and other physiological markers while also capturing psychological assessments throughout pregnancy [13]. Wearable devices, such as the Oura smart ring, provide objective data on heart rate, HRV, physical activity, and sleep patterns, allowing researchers to track and assess longitudinal health trends outside traditional clinical and laboratory settings [14, 15]. In conjunction, ecological momentary assessment (EMA) enables real-time self-reported data collection on emotional distress, capturing the psychological experiences of participants in their natural environment [16]. Integrating these approaches offers a more comprehensive assessment of both objective (physiological measures) and subjective (psychological states) factors that influence HRV during pregnancy.

Various factors contribute to fluctuations in HRV, including both physiological and psychological influences. Physiological stress, which is objectively measured through biomarkers such as HRV [10], contrasts with emotional distress, which refers to subjective experiences of psychological strain characterized by feelings of sadness, anxiety, and anger. However, emotional distress can also induce physiological responses that alter autonomic regulation and impact maternal and fetal health [17]. Specifically, emotional distress is associated with reduced HRV, suggesting a diminished capacity for physiological stress regulation [18, 19, 17]. On the other hand, physical activity (PA) has been consistently shown to positively influence HRV, improving autonomic function and facilitating physiological stress adaptation [20, 21].

However, the interaction between these two factors remains underexplored, particularly during pregnancy. While both physical activity and emotional distress independently influence HRV, it is unclear whether emotional distress moderates the relationship between physical activity and HRV. A moderator is a variable that alters the strength or direction of an association between an explanatory variable and the response variable through an interaction effect [22, 23]. In this case, emotional distress may act as a moderator, potentially altering the beneficial effects of physical activity on HRV. Understanding this interaction is especially relevant for pregnant populations, where both physiological and psychological factors contribute to maternal and fetal outcomes.

To address this knowledge gap, this study aims to conduct a comprehensive analysis incorporating both objective and subjective measures to examine their influence on HRV during pregnancy. Specifically, we aim to (1) examine how emotional distress and physical activity individually affect HRV, while adjusting for other factors such as age, pre-pregnancy body mass index (BMI), heart rate, and sleep parameters (deep and REM sleep), which are also known to influence HRV [24, 25, 26, 27, 28, 29, 30, 31, 32], and (2) examine whether emotional distress moderates the relationship between physical activity and HRV, i.e., whether the benefits of PA on HRV differ based on an individual’s level of emotional distress.

By integrating objective physiological data from wearable devices with subjective self-reports of emotional states, this study aims to enhance the understanding of the complex interactions between emotional distress, physical activity, and HRV in pregnant women. Ultimately, our findings could inform intervention strategies, encouraging physical activity as a potential means to regulate HRV during pregnancy and promote overall maternal and fetal health.

## 2 Materials and methods

### 2.1 Study design and sampling

The study used a longitudinal observational design. Following IRB (Institutional Review Board) approval, convenience sampling was employed to recruit participants by disseminating the study flyer through established local community partners working with underserved perinatal women in the Southern California region. The inclusion criteria were pregnant women aged 18-40 years, with a healthy singleton pregnancy, and access to a smartphone. The research coordinator explained the study procedure and obtained consent from participants using the IRB-approved Study Information Sheet. Once consent was obtained, the Oura ring wearable device was shipped to participants for data collection.

A total of 20 participants were eligible for the Oura ring data collection, but two dropped out due to family circumstances, leaving 18 who provided data on physiological metrics such as HRV and resting heart rate. More detailed information on these participants can be found in an earlier study [33]. Meanwhile, 17 participants provided EMA data through smartphone surveys, capturing self-reported emotional experiences in real time. Among them, only 13 had both Oura ring and EMA data. We retained 9 participants by selecting those with second trimester data and aligning the merged dataset, pairing each day’s Oura ring data with the previous day’s EMA data, to examine how daytime emotional states influenced subsequent nighttime physiological metrics.

### 2.2 Data collection

Self-reported demographic data, including maternal age, ethnicity, and pre-pregnancy body mass index (BMI), were collected using REDCap, a secure data collection platform [34]. The Oura ring, a waterproof wearable device equipped with multiple sensors, measured physiological signals through an optical pulse waveform detected from the participant’s finger, including heart rate variability (HRV), heart rate, and physical activity [15]. The ring automatically transferred data to an app on the participant’s smartphone via Bluetooth, delivering a daily summary.

Data on the participants’ various emotional states during pregnancy were also collected using daily ecological momentary assessment (EMA), a method used to capture real-time emotional states or moods by prompting participants to report their feelings during the day [16]. EMAs were delivered to participants daily over the course of the study at approximately noon via a mobile application. Responses were automatically saved and subsequently retrieved from the study dashboard for analysis. A description of the methods employed in developing the app-based platform can be found in an earlier study [14]. Specifically, the EMA prompts solicited information regarding participants’ current emotional states, encompassing both positive (healthy, cheerful, content, excited, safe) and negative (sad, lonely, angry, overwhelmed, stressed, tired, worried) feelings. By collecting data in real-time, EMA minimizes recall bias and captures fluctuations in emotional states as they occur, allowing for a better understanding of how emotions vary for an individual [35]. Physiological measures, such as HRV and resting heart rate, were collected overnight and reported at the start of each day, while emotional states were recorded later in the day via EMA.

### 2.3 Data preprocessing

In this study, we focused on the second trimester (clinically, 14–28 weeks gestation), a period characterized by significant physiological and cardiovascular changes [11, 12] but relatively greater emotional stability compared to the third trimester.

For our analysis, we combined the EMA data with physiological data from the Oura ring to jointly examine the relationship between emotional and physiological factors, as well as their interactions, on HRV during pregnancy. We aligned these data by pairing each day’s physiological measures with the previous day’s emotional state responses. This pairing allowed us to examine how daytime emotional states influenced subsequent physiological stress markers at night. After data preprocessing, we retained 9 subjects, with an average of 10 daily observations per subject (93 daily observations total).

In the daily EMA survey, a total of 12 emotions were evaluated, ranging from positive to negative emotions. Positive emotions are characterized by feelings of well-being and happiness, such as being cheerful, excited, healthy, content, and safe, while negative emotions involve states of distress and discomfort, including feeling tired, overwhelmed, stressed, worried, angry, lonely, or sad. The intensity of each emotion was assessed using a 5-point Likert scale [36] with values ranging from 0 (Not at all) to 4 (A lot). For example, participants were asked:

*”How sad are you feeling right now?”* Response options: 0 (Not at all), 1 (A little), 2 (Somewhat), 3 (Quite a bit), or 4 (A lot).

Figure 1 presents a heatmap of the pairwise correlations between the 12 emotion measurements, illustrating the overall relationships among different emotional states. The figure highlights the contrasting patterns between positive and negative emotions, offering insights into their co-occurrence and differentiation. In the heatmap, positive emotions (e.g., cheerful, excited, safe) are expected to show strong positive correlations with each other, indicating that individuals who frequently experience one positive emotion are likely to experience others in the same category. Conversely, negative emotions (e.g., tired, overwhelmed, sad) also exhibit strong positive correlations among themselves but weakly negative correlations with positive emotions.

**Figure 1:**
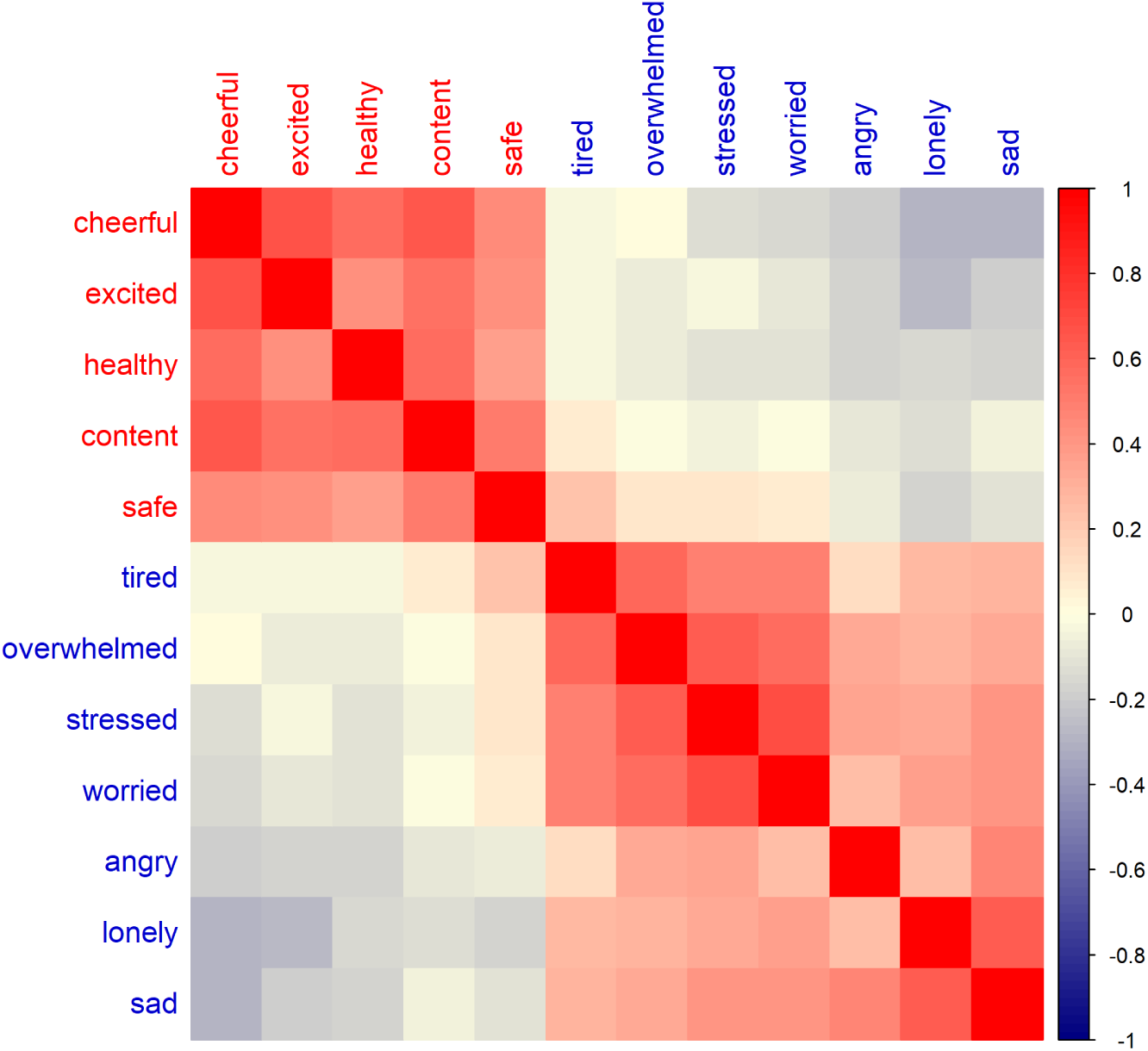
Heatmap of the pairwise correlations between the 12 emotions from the daily EMA data, where red indicates a strong positive correlation and blue indicates a strong negative correlation. Spearman correlation was used to measure the strength and direction of the monotonic relationship between the emotion variables.

In order to quantify emotional distress, we constructed an overall emotional distress (ED) score based on the 12 emotions measured through EMA. Before summing the 12 emotions, we redefined the five positive emotions (cheerful, excited, healthy, content, and safe) into their negated forms (e.g., not cheerful, not excited), ensuring that all items consistently reflected some level of negative emotional experience. To achieve this, we reverse-coded the Likert scale for the five positive emotions so that 0 became 4, 1 became 3, 2 remained 2, 3 became 1, and 4 became 0. This transformation allowed for a consistent interpretation where higher scores across all 12 emotions indicated greater emotional distress. The adjusted emotion ratings were then summed, resulting in an overall ED score, with higher values reflecting greater emotional distress. As shown in Figure 2, this composite score ranged from 6 to 40.

**Figure 2:**
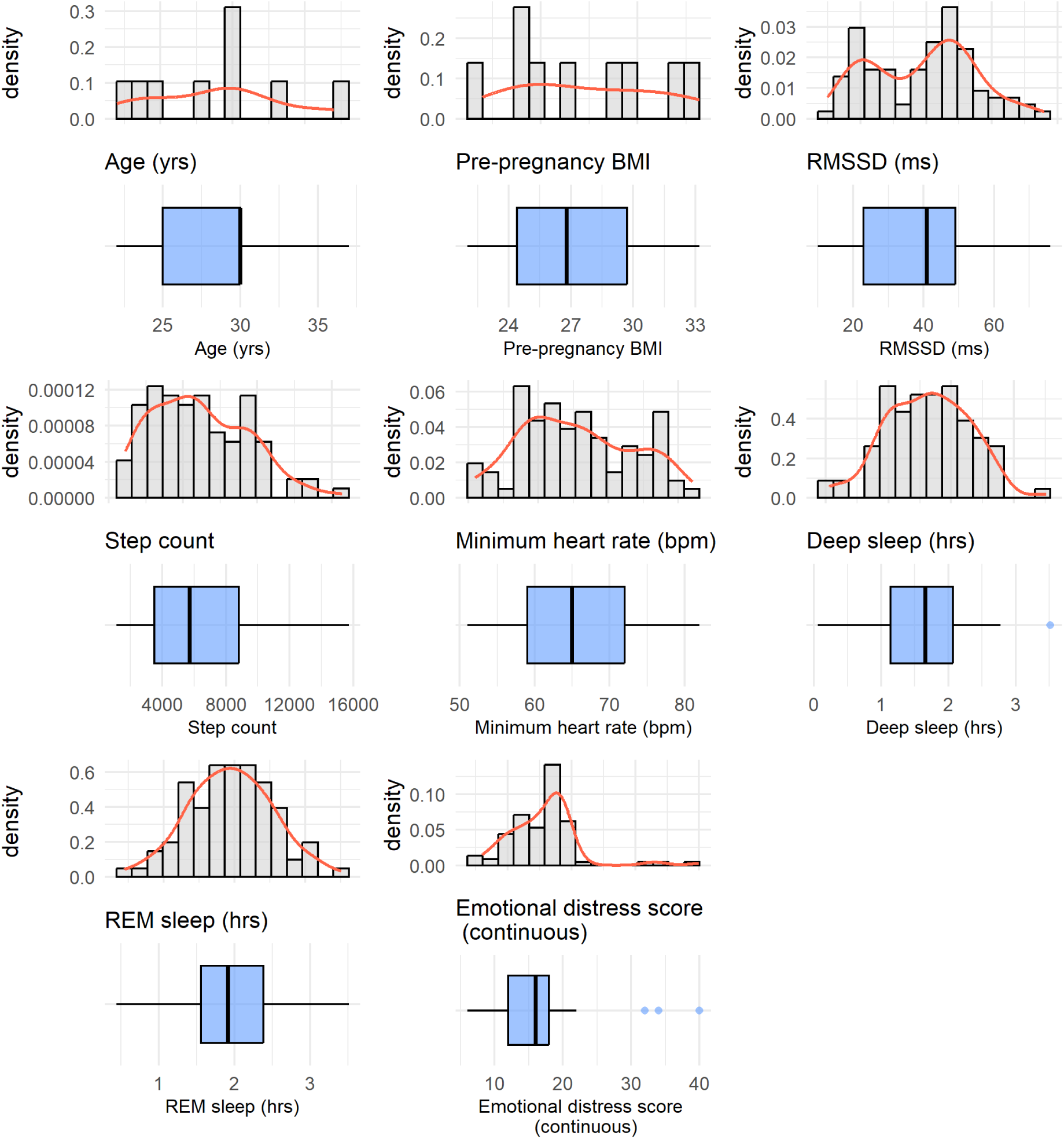
Distributions of variables considered in the model, presented as histograms and corresponding boxplots below. The histograms illustrate the estimated probability density of each variable, while the boxplots show the median, interquartile range, and potential outliers.

To enhance interpretability, we categorized the overall ED score into categorical levels of emotional distress using quantile-based cutoffs, dividing the scores into 3 groups (i.e., using 34th and 67th percentiles). Based on this approach, we defined the cutoffs as follows: scores 6–14 as low emotional distress, 15–18 as moderate emotional distress, and 19–40 as high emotional distress. This resulted in 39%, 41%, and 20% of observations in the low, moderate, and high emotional distress level, respectively. Due to the presence of many repeated values, it was difficult for the resulting groups to be more balanced.

### 2.4 Statistical analysis

#### 2.4.1 Primary outcome of interest

In this study, we examined potential factors that influence physiological stress among pregnant women. We used the root mean square of successive differences (RMSSD) as the objective measure for heart rate variability (HRV), which serves as an indicator of physiological stress. The RMSSD is calculated by taking the square root of the mean of the squares of successive differences between adjacent heartbeats. This measure is particularly useful for assessing the parasympathetic nervous system’s activity, which is responsible for promoting relaxation and recovery [37]. Higher RMSSD values indicate greater variability in heart rate, reflecting a more adaptable and resilient autonomic nervous system, which is often associated with lower physiological stress levels [9]. Conversely, lower RMSSD values suggest reduced variability and higher physiological stress levels, and a greater risk for cardiovascular and other health-related issues [38, 39]. In this paper, we use RMSSD and HRV interchangeably to refer to measures of autonomic regulation and cardiovascular health.

#### 2.4.2 Modeling

We examined the relationships between RMSSD, physiological and emotional factors (e.g., physical activity and emotional distress) using the generalized estimating equations (GEE) model, given the longitudinal nature of the data [40]. Due to repeated measurements from a limited number of subjects, subject-wise correlations on measured outcomes were expected to play a significant role in data analysis. Parameter estimations could be biased if within-subject correlations are ignored. The GEE approach, which accounts for the correlated structure of the data, produces more precise statistical analysis through a robust estimation of standard errors [40]. We specified an exchangeable correlation structure on the within-subject correlations.

Additionally, we used the log-transformed RMSSD as the response variable in the model. The analysis was performed using R software, and the GEE model was computed using the ‘geepack’ package. Moreover, we adjusted for five physiological and demographic variables in the model, including age, pre-pregnancy BMI, minimum resting heart rate, and deep and rapid eye movement (REM) sleep. These covariates were included based on their known associations with physiological stress, with prior research indicating their potential effects on HRV [24, 25, 26, 27, 28, 29, 30, 31, 32].

## 3 Results

Participants (*N* = 9) were primarily Hispanic women between the ages of 22 and 37 years, with an average age of 28.8 years (SD = 4.65) and an average pre-pregnancy BMI of 27.4 (SD = 3.84). Majority of the women were married (89%) and living with the father of the baby (89%). A third of the women were going through their first pregnancy. Summary statistics of the variables included in the GEE models are provided in Table 1 and their distributions are provided in Figure 2.

**Table 1:** Summary statistics for continuous variables considered in the model presented as mean *±* standard deviation and range, and for categorical variables as count (%) for *N* = 9 subjects with a total of 93 measurements.

| Variable | Mean ± SD or Count (%) | Range |
| --- | --- | --- |
| Age (yrs) | $28.8 \pm 4.7$ | (22, 37) |
| Pre-pregnancy BMI | $27.4 \pm 3.8$ | (22, 33.2) |
| RMSSD (ms) | $38.8 \pm 15.9$ | (10, 76) |
| Daily step count | $6,135 \pm 3,211$ | (1,125, 15,718) |
| Heart rate minimum (bpm) | $65.5 \pm 8.0$ | (51, 82) |
| Deep sleep (hrs) | $1.63 \pm 0.66$ | (0.06, 3.52) |
| REM sleep (hrs) | $1.94 \pm 0.60$ | (0.44, 3.51) |
| Daily emotional distress level: |  |  |
| Low | 36 (39%) | — |
| Moderate | 39 (41%) | — |
| High | 18 (20%) | — |

We first examined the individual effects of emotional distress and physical activity on HRV while adjusting for age, pre-pregnancy BMI, resting heart rate, and sleep parameters (deep and REM sleep). Since the response variable in our GEE models is the log-transformed RMSSD, all interpretations in this section are based on exponentiated coefficients. As shown in Table 2, holding all other variables constant, every additional 1,000 steps daily was associated with a 1.7% increase in RMSSD (*e*^0^.^000017^*^×^*^1000^ *≈* 1.017; *P*-value *≈* 0.008), suggesting that greater physical activity is associated with improved autonomic function.

**Table 2:** Model results examining the effects of emotional distress level (low, moderate, or high) and physical activity (step count) on HRV (log-transformed RMSSD), adjusting for covariates. Low emotional distress level was the reference level.

| | Estimate | Std. Err | Wald | $P$-value |
| --- | --- | --- | --- | --- |
| *(Intercept)* | 7.01 | 0.1990 | 1240.26 | < 0.001 *** |
| Age (yrs) | -0.0426 | 0.0045 | 91.18 | < 0.001 *** |
| Pre-pregnancy BMI | -0.0071 | 0.0023 | 9.61 | 0.0019 ** |
| **Step count** | 0.000017 | 0.0000063 | 6.96 | 0.0084 ** |
| Heart rate minimum (bpm) | -0.0346 | 0.0023 | 233.75 | < 0.001 *** |
| Deep sleep (hrs) | 0.0998 | 0.0294 | 11.50 | < 0.001 *** |
| REM sleep (hrs) | -0.0475 | 0.0162 | 8.65 | 0.0033 ** |
| **Moderate emotional distress** | -0.0017 | 0.0365 | 0.00 | 0.9628 |
| **High emotional distress** | 0.0046 | 0.0306 | 0.02 | 0.8818 |

*Significance codes:* 0 ‘\*\*\*’ 0.001 ‘\*\*’ 0.01 ‘\*’ 0.05 ‘.’ 0.1 ‘ ’ 1

Moreover, emotional distress levels showed distinct associations with HRV (see Table 2). As mentioned earlier, emotional distress was categorized into three levels: low, moderate, and high, with low emotional distress being the reference level in our model. Compared to low emotional distress, moderate emotional distress was associated with a 0.2% decrease in RMSSD, while high emotional distress was associated with a 0.5% increase in RMSSD. However, neither of these associations was statistically significant, indicating that emotional distress alone may not have a strong main effect on HRV.

Therefore, we included an interaction term between step count (physical activity) and emotional distress levels to examine whether the effect of physical activity on HRV varies by emotional distress level. The results in Table 3 revealed a statistically significant interaction between step count and high emotional distress (*P*-value *<* 0.001), suggesting that the relationship between physical activity and HRV depends on emotional distress level. Specifically, for subjects experiencing high emotional distress, each additional 1,000 steps was associated with a 3.5% increase in RMSSD compared to those experiencing low emotional distress. In contrast, the interaction between step count and moderate emotional distress was not statistically significant, and when emotional distress was low, step count had no significant effect on HRV. Thus, after adding the interaction term, the main effect of physical activity on HRV was no longer statistically significant, indicating that the influence of physical activity on HRV is not uniform across all emotional distress levels. Instead, its effect is heterogeneous, varying based on emotional distress levels. Our findings also suggest that the positive impact of physical activity on improving HRV is most pronounced on days of high emotional distress, compared to days of low or moderate emotional distress.

**Table 3:** Model results examining the moderating effect of emotional distress level (low, moderate, or high) on the relationship between physical activity (step count) and HRV (log-transformed RMSSD), adjusting for covariates. Low emotional distress level was the reference level.

|  | Estimate | Std. Err | Wald | *P*-value |
| --- | --- | --- | --- | --- |
| *(Intercept)* | 7.12 | 0.2390 | 889.20 | < 0.001 *** |
| Age (yrs) | -0.0412 | 0.0050 | 67.38 | < 0.001 *** |
| Pre-pregnancy BMI | -0.0092 | 0.0027 | 11.37 | < 0.001 *** |
| Step count | 0.0000052 | 0.0000048 | 1.17 | 0.2795 |
| Heart rate minimum (bpm) | -0.0349 | 0.0025 | 194.70 | < 0.001 *** |
| Deep sleep (hrs) | 0.1030 | 0.0260 | 15.59 | < 0.001 *** |
| REM sleep (hrs) | -0.0527 | 0.0142 | 13.75 | < 0.001 *** |
| Moderate emotional distress | -0.0336 | 0.0465 | 0.52 | 0.4700 |
| **High emotional distress** | -0.2060 | 0.0335 | 37.78 | < 0.001 *** |
| Step count × Moderate emotional distress | 0.0000062 | 0.0000068 | 0.84 | 0.3606 |
| **Step count × High emotional distress** | 0.000035 | 0.0000042 | 68.34 | < 0.001 *** |

*Significance codes:* 0 ‘\*\*\*’ 0.001 ‘\*\*’ 0.01 ‘\*’ 0.05 ‘.’ 0.1 ‘ ’ 1

Figure 3 visually illustrates the heterogeneous effects of daily step count on HRV across different emotional distress levels, according to the model presented in Table 3. The figure depicts the predicted RMSSD as a function of daily step count at different levels of emotional distress for a pregnant woman with average values for age, pre-pregnancy BMI, resting heart rate minimum, and sleep measures (deep and REM), as summarized in Table 1. This visualization highlights how the relationship between physical activity and HRV differs depending on emotional distress levels while holding other covariates constant.

**Figure 3:**
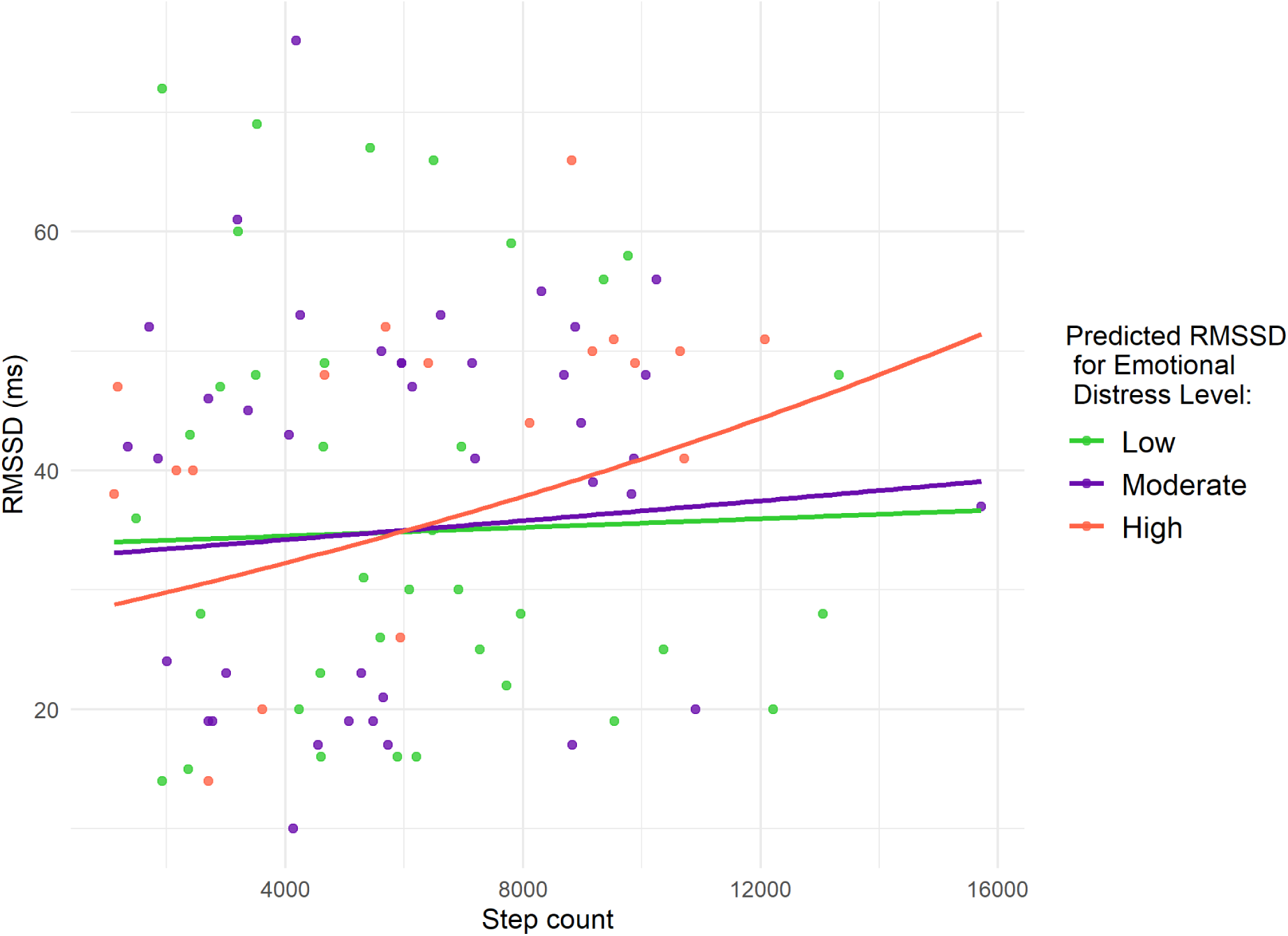
Predicted RMSSD as a function of daily step count at different levels of emotional distress for a pregnant woman with average age, pre-pregnancy BMI, minimum heart rate, and deep and REM sleep. Points represent observed daily measurements for the nine pregnant women, color-coded by emotional distress level.

Additionally, the analysis indicated that high emotional distress was significantly associated with lower HRV compared to low emotional distress. Specifically, on days with no physical activity (zero daily steps), high emotional distress corresponded to a 19% reduction in RMSSD compared to low emotional distress days. In contrast, moderate emotional distress did not show a significant association with HRV relative to low emotional distress under these conditions.

Several covariates were also significantly associated with RMSSD, and similar effects were observed for both models (with and without the interaction term). Older age was significantly associated with lower RMSSD, with each additional year of age corresponding to a 4.0% decrease in RMSSD. Pre-pregnancy BMI was negatively associated with RMSSD, with each unit increase in BMI corresponding to a 0.9% decrease in RMSSD. Higher resting heart rate was strongly associated with lower HRV, with each 1 bpm increase in heart rate corresponding to a 3.4% decrease in RMSSD. Regarding sleep, REM sleep was negatively associated with RMSSD, with each additional hour of REM sleep corresponding to a 5.1% decrease in RMSSD. In contrast, deep sleep was positively associated with RMSSD, where each additional hour of deep sleep corresponded to a 10.8% increase in RMSSD. These observed effects were statistically significant across all models.

## 4 Discussion

Our findings align with prior research demonstrating that emotional distress exacerbates reductions in HRV during pregnancy. A study reported significant declines in HRV during the first and second trimesters, with more pronounced reductions in pregnant women experiencing higher distress levels [17]. Consistent with this, our results showed that high emotional distress was associated with lower HRV, particularly on less active days, reinforcing the detrimental impact of distress on autonomic function. However, such an association was not observed for low or moderate emotional distress levels.

Furthermore, although existing research generally supports PA’s beneficial effects on HRV and emotional distress, our findings emphasize that PA provides a more pronounced protective effect on HRV during high distress days. This aligns with a study showing that prenatal yoga, a form of PA, increases HRV and alleviates emotional distress, suggesting a regulatory role of PA in autonomic function [21]. Similarly, another study found that regular PA is associated with lower resting heart rate and higher HRV during pregnancy, further supporting PA’s role in cardiovascular adaptation [20]. Additionally, a study suggested that PA may be protective against emotional distress during pregnancy, reinforcing its potential role in buffering autonomic dysfunction [41]. Thus, our study builds upon this body of work by demonstrating that PA can mitigate the adverse effects of emotional distress, particularly on days of elevated emotional distress.

In summary, our findings highlight the complex interplay between emotional distress, PA, and HRV during pregnancy, with emotional distress moderating the relationship between PA and HRV. Notably, the buffering effect of PA on HRV was most pronounced on days of high emotional distress, where each additional step had the strongest positive impact on improving HRV. However, the beneficial effects of PA on HRV were not observed on days of low or moderate emotional distress. This suggests that PA may serve as a powerful compensatory mechanism to mitigate the adverse effects of emotional distress on autonomic function [42].

A notable strength of this study is the use of real-time emotional data captured through ecological momentary assessment, which provides a more nuanced understanding of emotional fluctuations and their impact on physiological stress. Unlike traditional retrospective self-report measures, EMA offers a dynamic, real-time snapshot of emotional states, minimizing recall bias and providing more accurate insights into the relationship between emotions and physiological stress [16]. The integration of physiological data from the Oura ring, which offers objective measurements of HRV and physical activity, enhances our analysis by enabling us to explore the interaction between emotional distress and physical activity on autonomic function in a real-world context.

### 4.1 Implications

Our findings have important implications for intervention strategies aimed at improving maternal health. Given that emotional distress moderates the relationship between PA and HRV, promoting movement-based interventions such as walking programs or structured prenatal exercise could be particularly beneficial for pregnant women experiencing high emotional distress. Encouraging pregnant individuals to engage in daily PA, especially on days of high emotional distress, may serve as a practical and accessible strategy to enhance autonomic function and overall well-being.

Furthermore, the integration of mHealth technologies, such as wearable devices, enables real-time monitoring of HRV and PA patterns, allowing for personalized recommendations tailored to an individual’s emotional and physiological state. Given the increasing prevalence of mHealth tools in prenatal care, future research should explore how digital interventions can optimize PA engagement to improve maternal cardiovascular health, especially for individuals experiencing high levels of emotional distress.

### 4.2 Limitations

The study has some limitations to consider when interpreting the results. The sample size is small with only 9 pregnant women, and convenience sampling was used to recruit participants. As a result, the findings may not be broadly generalizable. The high proportion of Hispanic participants in our sample limits the generalizability of the findings to broader populations. Thus, future research with a larger sample size of pregnant women from diverse ethnic backgrounds is needed to confirm these results.

Additionally, the use of objective measures from the Oura ring and subjective measures from EMA data may introduce biases. Participants have access to their Oura ring data through their smartphone app, which could potentially influence their behavior and compromise data accuracy. The subjective nature of the emotional data collected through EMA also carries the potential for bias, as emotions may be influenced by factors beyond the study’s scope. In particular, although our study retained physiological biosignals and real-time evaluations of emotional states, a comprehensive understanding of physiological stress should extend beyond these measures to also include contextual information, such as the environment, life circumstances, and coping strategies [43, 44, 45, 46]. Specifically, EMA surveys were limited to emotional evaluations that did not discuss experiences or environmental details, which can provide additional information on HRV evaluation during pregnancy.

## 5 Conclusion

Our findings emphasize the heterogeneous impact of emotional distress on the relationship between physical activity and heart rate variability. Although emotional distress alone did not show a significant direct association with HRV, its interaction with physical activity revealed a significant moderating effect. On days when pregnant women experienced high emotional distress, increased physical activity was associated with a greater improvement in HRV, suggesting that being more active can help mitigate the adverse effects of emotional distress on autonomic function. This protective effect was not observed on days with low or moderate emotional distress, indicating that physical activity may be particularly beneficial on days with increased emotional distress. These results highlight the importance of being active in maintaining autonomic regulation and cardiovascular health, particularly in emotionally demanding settings. Future research should explore the mechanisms underlying this interaction and assess whether targeted interventions promoting physical activity could serve as a strategy to protect against the negative physiological effects of emotional distress during pregnancy.

## Data Availability

The mobile health data for pregnant women are collected by co-author Yuqing Guo. We will make the de-identified Oura ring data available to other researchers as requested following the completion of the present study. Access the data needs to follow IRB regulations.

## Acknowledgments

This research was supported by National Cancer Institute SCH: Individualized Learning and Prediction for Heterogeneous Multimodal Data from Wearable Devices (R01: CA297869), National Science Foundation Collaborative Research: Integrative Heterogeneous Learning for Intensive Complex Longitudinal Data (DMS: 2210640), and Smart and Connected Communities (DCNS: 1831918).

## Author Contributions

**Conceptualization:** Jenifer Rim, Qi Xu, Xiwei Tang, Annie Qu, Yuqing Guo.

**Data curation:** Jenifer Rim, Tamara Jimah.

**Formal analysis:** Jenifer Rim, Qi Xu, Xiwei Tang.

**Funding acquisition:** Annie Qu, Yuqing Guo.

**Investigation:** Jenifer Rim, Annie Qu, Yuqing Guo, Tamara Jimah.

**Methodology:** Jenifer Rim, Annie Qu, Yuqing Guo.

**Project administration:** Jenifer Rim, Annie Qu.

**Software:** Jenifer Rim.

**Supervision:** Annie Qu, Yuqing Guo, Tamara Jimah.

**Validation:** Jenifer Rim, Annie Qu.

**Visualization:** Jenifer Rim, Annie Qu.

**Writing - original draft:** Jenifer Rim.

**Writing - review & editing:** Jenifer Rim, Qi Xu, Xiwei Tang, Tamara Jimah, Yuqing Guo, Annie Qu.

